# Troponin antibodies and macrotroponin: a common cause of troponin I elevation in elite endurance athletes

**DOI:** 10.1101/2025.11.16.25340346

**Authors:** Stephanie Rowe, Kalayani McKubre, Kristel Janssens, Amy Mitchell, Oscar Cullen, Luke Spencer, Youri Bekhuis, Paolo D’Ambrosio, Kaitlin Newcomb, Elizabeth Paratz, Rik Willems, Hein Heidbuchel, Guido Claessen, Christina Trambas, Andre La Gerche

## Abstract

**Background and Aims:** Elevations in cardiac troponin I (cTnI), a marker of myocardial injury, have been frequently observed in athletes after intense exercise. cTnI autoantibodies (cTnI Ab) and macrotroponin complexes can confound assessment, and we hypothesised that they may explain the paradox of repeated cTnI elevations in athletes.

**Methods:** Athletes and non-athlete controls underwent high-sensitivity cTnI testing, cardiac magnetic resonance imaging (CMR), 24-hour ambulatory electrocardiographic monitoring and VO_2_peak assessment. Athletes with cTnI values >99^th^ percentile sex-specific upper reference limit were assessed for cTnI Ab using polyethylene glycol (PEG) precipitation and troponin T (TnT)/TnI discordance.

**Results:** Overall, 456 participants were evaluated (387 athletes and 69 non-athletes). cTnI elevations were observed in 39 (10.1%) athletes and no (0%) control participants (p=0.002). Athletes with cTnI elevation demonstrated higher VO_2_peak and larger biventricular dimensions compared to athletes without cTnI elevations (p<0.001). Of the athletes with cTnI elevations, 31 of 39 (79%) had evidence of cTnI Ab; 26 (66.7%) met both criteria for cTnI Ab (low PEG recovery and normal cTnT), a further 5 (12.8%) met one criteria (low PEG recovery but slightly elevated cTnT out of keeping with cTnI elevation [median TnT/TnI=3.7%]) and the remaining 8 athletes met neither (high PEG recovery and cTnT elevation [median TnT/TnI=81.2%]). Four of these 8 athletes with TnI elevation but without cTnI Ab had evidence of possible myocardial injury on CMR.

**Conclusions:** cTnI elevations are commonly found in highly-trained endurance athletes, especially the fittest athletes with greatest cardiac remodelling, but are due to antibodies in a majority.

**Graphic Abstract:** *Key Question:*

- What is the prevalence of cardiac troponin I (cTnI) elevation in highly-trained endurance athletes outside of competition and what proportion are due to cTnI antibodies?
- How do elevated cTnI measures relate to exercise-induced cardiac remodelling and VO_2_peak?

*Key Finding:*

- 1 in 10 endurance athletes had elevated cTnI with 80% due to cTnI antibodies as opposed to ‘true’ cTnI elevations.
- cTnI elevation and antibodies were found in the fittest athletes with the greatest cardiac remodelling on cardiac magnetic resonance imaging.

*Take-home message:*

- cTnI antibodies are highly prevalent in endurance athletes and are not associated with myocardial damage.
- Awareness of cTnI antibodies and appropriate testing will help avoid intensive investigation and misdiagnoses. 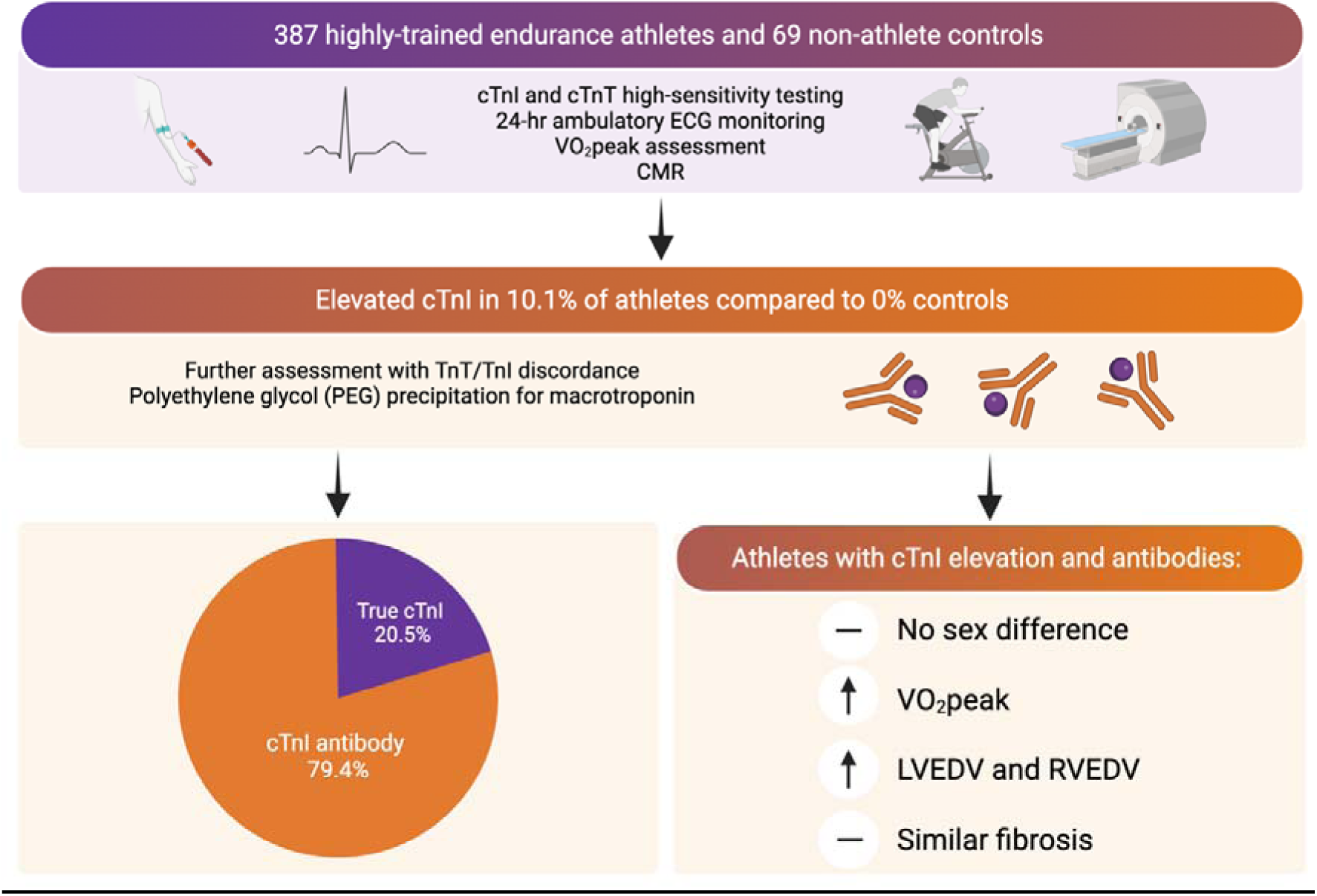

**Non-standard abbreviations/acronyms:**

cTnI = cardiac troponin I cTnT = cardiac troponin T
PEG = polyethylene glycol precipitation Ab = antibody
LGE = late gadolinium enhancement
NSVT = non-sustained ventricular tachycardia

## Introduction

High-sensitivity cardiac troponin assays are central to the diagnosis of myocardial infarction and myocarditis, conditions of particular concern in athletes because of their association with sudden cardiac death and the implications for safe return to play^1,2^. Yet, paradoxical troponin elevations are frequently observed after endurance exercise in otherwise healthy athletes and often these elevations would be considered clinically significant with more than half exceeding thresholds deemed consistent with myocardial damage^3–6^. Although post-exercise elevations in cardiac troponin are well described, the prevalence of cardiac troponin elevations in non-exercise related settings has seldom been interrogated. In clinical practice, such findings raise concern for myocarditis, myocardial infarction or pulmonary embolism and trigger intensive investigation^7,8^.

Despite decades of research, the biological basis of exercise-related troponin elevations remain uncertain. Proposed mechanisms include changes to cardiomyocyte membrane permeability, apoptosis or necrosis^9^. However, none of these mechanisms are proven, and there seems to be an unexplained paradox in which repeated bouts of myocardial injury could co-exist with athletic cardiac remodelling. Regular endurance training promotes an increase in cardiac volumes and mass that is proportional to increases in cardiorespiratory fitness^10,11^. However, there is consensus that there is limited capacity for cardiac muscle regeneration^12^. Thus, the concept of damage-induced regeneration and super-compensation as an explanation for athletic cardiac hypertrophy seems invalid and the observation of recurrent troponin elevations remains unexplained.

The troponin complex is formed by cardiac troponin I (cTnI), T (cTnT) and C and is essential for cardiac muscle contractility^13^. Both cTnI and cTnT are mainly localized to the sarcomere of cardiomyocytes and therefore immunoassays to detect and quantify circulating cTnI or cTnT are widely used to detect myocardial injury. Current high-sensitivity assays utilize capture and detection antibodies to detect circulating cardiac troponin. However, assay performance can be compromised by endogenous antibodies that react with animal immunoglobulins used in assays (heterophile antibodies) and by macrocomplexes comprising clumps of antibodies bound to troponin termed ‘macrotroponin’^14,15^. Autoantibodies against cTnI have been detected in up to 20% of healthy individuals^16,17^ and at even greater prevalence in individuals with cardiomyopathy^17^, although significant assay interference resulting in ‘false positive’ troponin elevations are less common and are a seldom considered confounder in clinical practice^14^.

We hypothesized that cTnI autoantibodies and subsequent macrotroponin formation were a potential cause of cTnI assay interference in athletes. To address this, we examined their prevalence and associations with cTnI levels, fitness, cardiac remodelling and myocardial scar in a large cohort of highly-trained endurance athletes and non-athlete controls.

## Methods

### Study Design and Population

This study involved participants enrolled in either the Pro@Heart study (ACTRN12618000716268) or the ProAFHeart study (ACTRN12618000711213) from medical and research facilities in Melbourne, Australia (Baker Heart and Diabetes Institute; St Vincent’s Institute of Medical Research). Both studies shared the same protocol which has been described in detail elsewhere^18^. Enrolled participants were current or former elite endurance-trained athletes (currently or previously competing in endurance sports at national or international level for at least two years) or non-athletic control subjects (participated in < 3 hours per week of organized sport). All participants underwent assessment between July 2015 and September 2024. Clinical and lifestyle information was collected by questionnaire. Participants were included in this analysis if they were ≥ 16 years old and had undergone assessment of high-sensitivity cardiac Troponin I at enrolment (cTnI). Participants were excluded if they had a known history of cardiomyopathy or moderate or greater valvular heart disease. Ethical approval was granted by the Alfred Hospital Human Research Ethics Committees.

### Assessment of troponin and antibodies

Samples were collected from participants in a rested state outside of a competitive setting and athletes were encouraged to avoid vigorous training in the days preceding testing. After venipuncture, cTnI was measured with the Abbott Architect high-sensitivity assay and then samples were stored at -80 degrees Celsius. cTnT was later analysed using Roche Elecsys high-sensitivity assay. Troponin I measures were considered elevated if the value was above the 99^th^ percentile upper reference limit (URL) for sex (cTnI ≥26ng/L for males and ≥16ng/L for females). Troponin T measures were considered elevated if values were ≥15ng/L. Samples of participants with an elevated cTnI at enrolment were further analysed with (i) repeat testing for cTnI to confirm elevation (ii) polyethylene glycol (PEG) precipitation to assess for the presence of cTnI antibodies and macrotroponin (iii) TnT/TnI discordance. A threshold of <25% recovery of the initial cTnI measurement following PEG precipitation was used to assess for the possible presence of anti-troponin antibodies (cTnI Ab). This threshold was derived in our laboratory via RefineR reference interval determination based on PEG precipitation of >1000 samples with troponin elevation (not shown) and is similar to thresholds published elsewhere^19,20^. Discordance between cTnT and cTnI (TnT/TnI) was assessed using the cTnT measure divided by the cTnI for a participant and presented as a percentage.

### Cardiopulmonary Exercise Testing (CPET)

Peak oxygen consumption (VO_2_peak) was determined from CPET performed on a cycle ergometer (LODE Excalibur Sport, Groningen, The Netherlands) using the highest 30 second rolling average of values from breath-by-breath measures using a calibrated metabolic cart (JaegerTM VyntusTM CPX, Vyaire Medical, Mettawa, IL) with heart rate (HR) and rhythm assessed from a 12-lead ECG, The percentage of predicted VO_2_peak was derived using the FRIEND equation^21^.

### CMR Imaging

CMR was performed using a Prisma 3.0T or Sola 1.5T (Siemens Healthineers, Erlangen, Germany). Steady-state free precession (SSFP) short-axis and 4-chamber cine imaging was obtained during breath-hold to quantify biventricular function and volumes and left ventricular cardiac mass. Left ventricular mass was quantified by epicardial and endocardial contouring on short-axis slices at end-diastole, excluding papillary muscles and trabeculae. Native and post contrast T1 mapping was performed using the Shortened Modified Look-Locker Inversion recovery (ShMOLLI) method sequence. Haemoglobin measures from the same day were integrated to calculate extracellular volume (ECV). Myocardial fibrosis was assessed by late-gadolinium enhancement (LGE) imaging on breath hold phase-sensitive inversion recovery sequences 10 minutes after administration of gadolinium-DTPA after optimization of inversion recovery time to ensure good myocardial nulling. Analysis was performed by experienced investigators using Circle Cardiovascular Imaging (Calgary, Canada).

### Holter Monitor

Ambulatory 24-hour electrocardiographic Holter monitoring was performed using a PocketECG Holter (MEDICalgorithmics, Warsaw, Poland). Non-sustained ventricular tachycardia (NSVT) was defined as >3 consecutive ventricular beats at a rate of >100/min and lasting <30s.

### Statistical Analyses

Continuous variables are presented as means ± standard deviation (SD) or medians [25^th^ - 75^th^ percentile] according to normal distribution status. Categorical variables were expressed as frequencies and percentages. A Kruskal-Wallis or One Way ANOVA test was run to determine if there were differences in dependent variables between three groups of participants (non-athlete controls, athletes with normal cTnI and athletes with elevated cTnI). Distributions of the dependent variables were all assessed visually. Chi-square testing or the Fisher’ s Exact test was used for categorical variables. Pairwise comparisons were performed using Dunn’s (1964) procedure with a Bonferroni correction for multiple comparison. Statistical significance was accepted at a 2-sided p <0.05. Statistical analyses were performed using STATA v17.0 (STATACorp, Texas, United States of America).

## Results

### Study Cohort

Overall, 456 participants were evaluated (387 highly-trained endurance athletes [74% male] and 69 non-athlete participants [70% male]). Measures of cTnI were available in all participants and cTnT measures were available in 91.4%. Of the athletes, 39 (10.1%) had a cTnI measure elevated above the 99^th^ percentile sex-specific cut-off (*Table S1*). No control participants had an elevated cTnI measure (p < 0.001). Most cTnI elevations were modestly elevated (< 100 ng/L, *Table S1*) but in 6 subjects the cTnI exceeded 100 ng/L (15.4% of those with a cTnI elevation and 1.5% of the total athlete population). *Table 1* summarizes the baseline characteristics and CPET results based on cTnI status. *Table S2* summarizes these characteristics based on cTnI and cTnI antibody status. Compared to athletes (with or without cTnI elevation), control participants had lower cTnI and achieved a lower workload and VO_2_peak during exercise testing (p <0.001). The median cTnI was <2ng/L for non-athletes (limit of quantitation of cTnI is 2ng/L), 3 [2-6]ng/L for athletes with normal cTnI and 53 [34 – 84] ng/L for athletes with an elevated cTnI (p <0.001). Athletes with an elevated cTnI achieved a higher workload (416 ± 70 watts vs 364 ± 92 watts, p = 0.002), and higher VO_2_peak (60.0 [50.1 – 69.1] ml/kg/min vs 47.8 [37.1 – 58.6] ml/kg/min, p <0.001) compared to athletes without an elevated cTnI.

**Table 1.** Baseline characteristics, cardiopulmonary exercise testing and Holter monitoring by troponin status

|  | Non-athletes<br>(n=69) | Athletes with<br>normal cTnI<br>(n=348) | Athletes with<br>elevated cTnI<br>(n=39) | P value |
| --- | --- | --- | --- | --- |
| Age (yrs) | 23 [21 – 34] | 44 [23-60] * | 23 [19 – 37] ‡ | <0.001 |
| Male Sex, n (%) | 48 (69.6) | 257 (73.9) | 29 (74.4) | 0.753 |
| BMI (kg/m<sup>2</sup>) | 24.3 [22.2 – 26.9] | 23.7 [22.1 -25.5] | 21.8 [20.8 – 23.7] †‡ | 0.001 |
| Sport, n(%) | - | Athletics = 28 (8.0)<br>Cycling = 87 (25.0)<br>Rowing = 162 (46.6)<br>Other = 71 (20.4) | Athletics = 2 (5.1)<br>Cycling = 21 (53.9)<br>Rowing = 8 (20.5)<br>Other = 8 (20.5) | - |
| Haemoglobin (g/L) | 146 ± 11.2 | 144 ± 9.9 | 142 ± 9.1 | 0.125 |
| Creatinine (µmol/L) | 72 [64 – 82] | 73 [68 – 81] | 70 [65 – 76] ‡ | 0.039 |
| cTnI (ng/L) | < 2ng/L | 3 [2 – 6] * | 53 [34 – 84] †‡ | <0.001 |
| cTnT (ng/L)<sup>§</sup> | 4.4 [3.2 – 5.9] | 6.4 [4.9 – 8.8]* | 9.6 [7.0 – 24.0]†‡ | <0.001 |
| Cardiopulmonary Exercise Testing |  |  |  |  |
| Rest HR (bpm) | 75 [67 – 87] | 55 [48 – 64]* | 50 [46 – 56] † | <0.001 |
| Rest SBP (mmHg) | 134 [121 – 144] | 141 [130 – 150] * | 147 [133– 153] † | 0.002 |
| Rest DBP (mmHg) | 80 [71 – 88] | 85 [77 – 94] * | 84 [71 – 91] | 0.003 |
| Peak HR (bpm) | 190 [183 – 195] | 176 [165 – 188] * | 183 [174 – 192] | <0.001 |
| Peak SBP (mmHg) | 192 [173 – 212] | 213 [194 – 232] * | 215 [204 – 237] † | <0.001 |
| Peak DBP (mmHg) | 81 [75 – 88] | 85 [76 – 94] * | 82 [72 – 90] | 0.008 |
| Peak Power (W) | 266 ± 79 | 364 ± 92* | 416 ± 70 †‡ | <0.001 |
| VO<sub>2</sub>peak (ml/min) | 2919 [2302 - 3482] | 3669 [2851 – 4437] * | 4118 [3691 – 4831] †‡ | <0.001 |
| VO<sub>2</sub>peak (ml/kg/min) | 38.2 [31.6 - 44.1] | 47.8 [37.1 – 58.6] * | 60.0 [50.1 – 69.1] †‡ | 0.001 |
| VO<sub>2</sub>peak (% predicted) | 89.7 [76.8 – 98.0] | 126.5 [112.0 – 139.3] * | 133.3 [124.4 – 145.8]†‡ | <0.001 |
| Holter Monitoring<sup>#</sup> |  |  |  |  |
| Any PVC, n (%)<sup>#</sup> | 28 (41) | 232 (68)* | 25 (64)† | <0.001 |
| PVC ≥ 100, n (%) | 6 (8.8) | 63 (18.5) | 10 (25.6) | 0.086 |
| PVC ≥ 500, n (%) | 1 (1.5) | 26 (7.6) | 6 (15.4) † | 0.020 |
| NSVT, n (%) | 0 (0) | 16 (4.7%) | 3 (7.7%) | 0.058 |

Troponin T elevation was present in 7.7% (n = 30) of the athletes compared to zero controls (p = 0.009). Of the athletes with normal cTnI, 17 had elevated cTnT (4.9%) with a median value of 16.6ng/L (normal <15ng/L) and a range of 15.2ng/L–24.3ng/L. In comparison, 33.3% of athletes with elevated cTnI had a concurrent elevation in cTnT (median 29.5ng/L, range 17.5 – 57.8ng/L).

After comprehensive evaluation that included exercise testing, CMR and Holter monitoring as part of the study protocol – no athletes were diagnosed with cardiomyopathy, myocarditis or ischemic myocardial damage.

### Prevalence of Suspected Troponin I antibody in athletes with Troponin I elevation

Of the 39 athletes with cTnI elevation, 79.5% (n =31) had PEG recovery fractions suggestive of cTnI Ab (*Figure 1 and 2*). The majority of these athletes had normal cTnT results (80.6%). Five athletes suspected of having cTnI Ab had mild cTnT elevation with discordance between cTnT and cTnI levels (TnT/TnI =3.7%, *Table S3*). The two highest cTnI values were 4769ng/L and 1671ng/L. Both athletes had PEG recovery fractions <25% suggestive of cTnI Ab. In both cases, cTnT was also elevated, but out of keeping with the level of cTnI (TnT 26.4ng/L [0.5% of cTnI measure] and 44.3ng/L [2.7% of cTnI], respectively).

**Figure 1.**
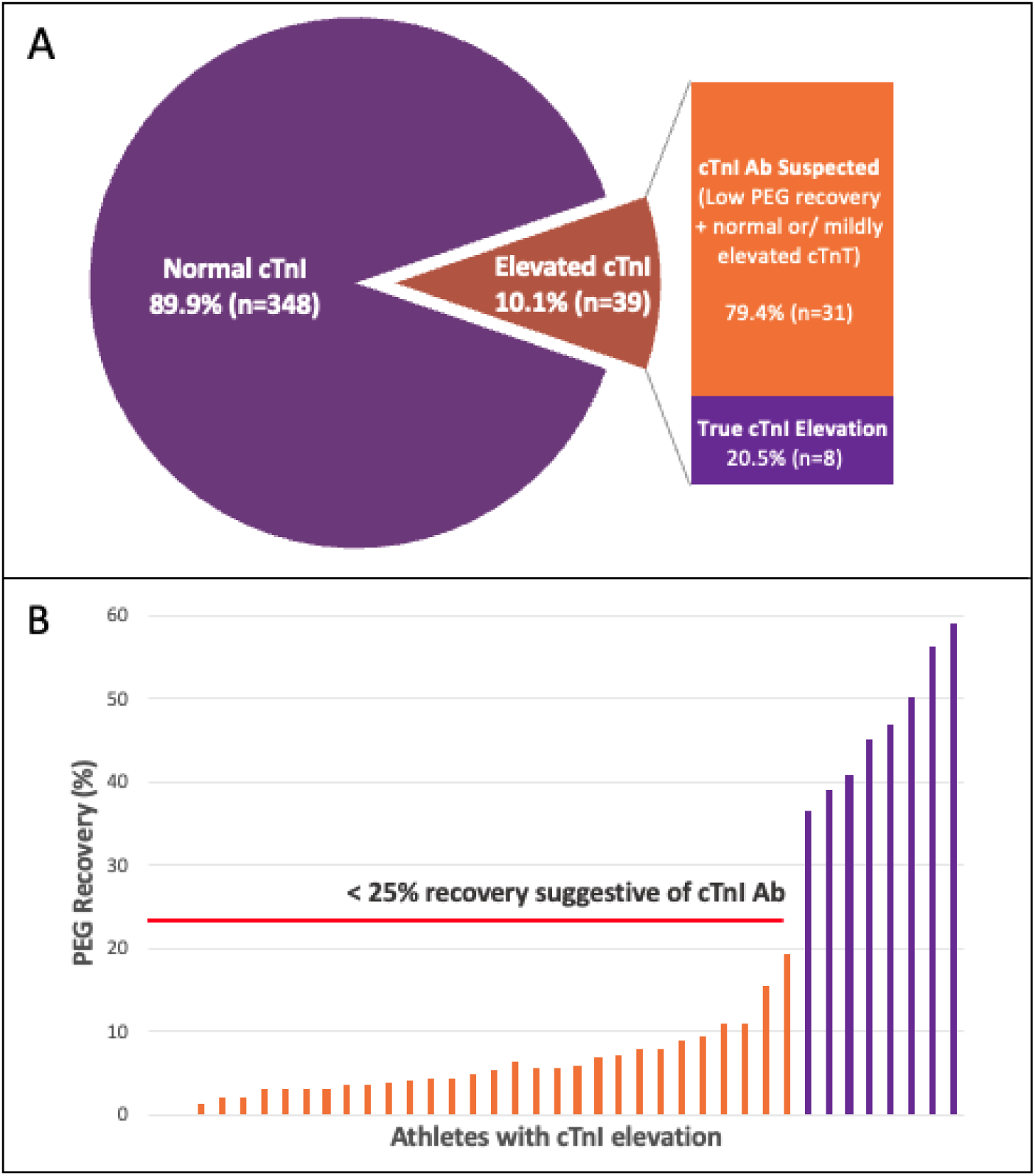
*Prevalence of cTnI Ab in athletes with elevated cTnI measures.* The bar of pie chart (Panel A) demonstrates the prevalence of cTnI elevation and the proportion of athletes with suspected cTnI Ab. Nearly 80% of athletes with elevated cTnI had suspected cTnI Ab. The bar chart (Panel B) demonstrates the percentage recovery of the initial cTnI measurement following precipitation with polyethylene glycol (PEG), with low recovery < 25% consistent with the presence of cTnI Ab.

**Figure 2.**
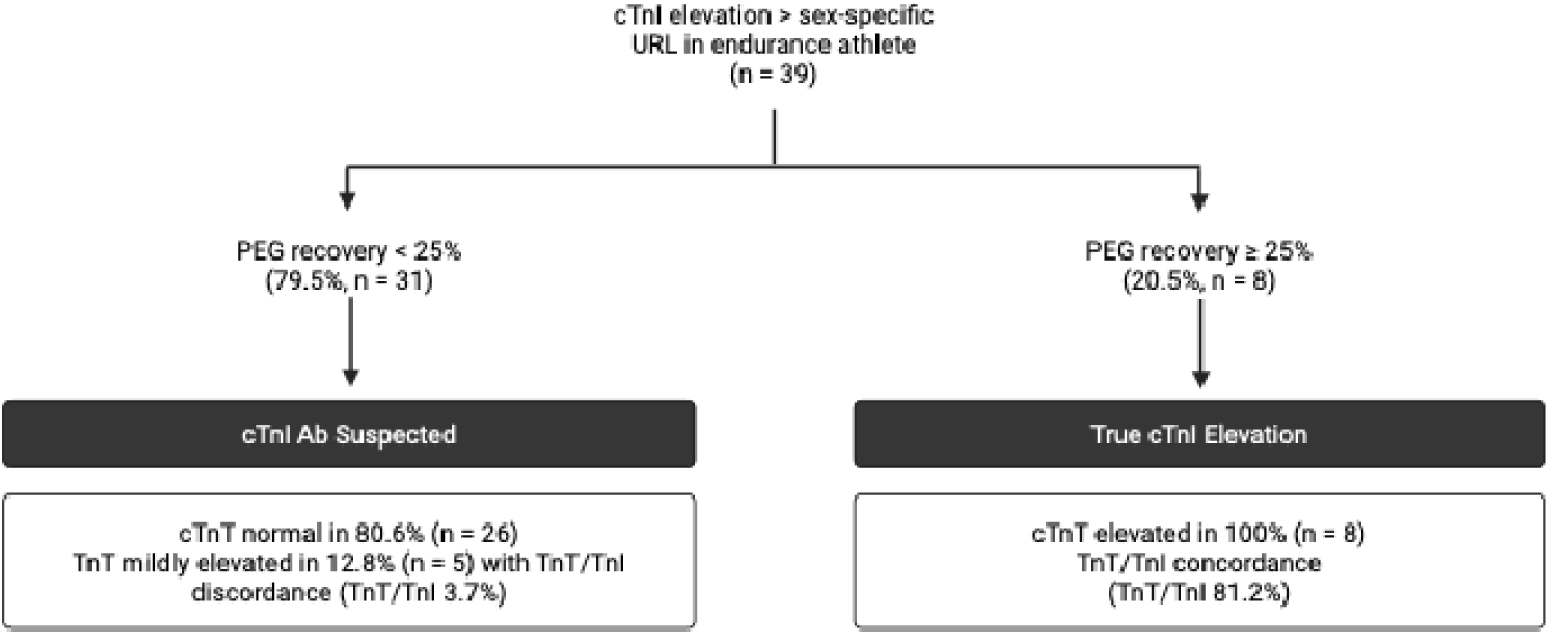
*Prevalence of cTnT elevation in athletes with and without cTnI Ab.* The flow chart shows prevalence of cTnT elevation in each subgroup and the concordance between cTnT and cTnI results. Created with BioRender.com.

Overall, 8 athletes (2%) had PEG recovery fractions > 25% suggesting true cTnI elevation – all of these athletes had cTnT elevation with high concordance between TnT/TnI measures (81.2%). Four of these athletes had evidence of possible myocardial pathology on CMR (2 with mid-wall late LGE and small pericardial effusions, and 2 with small pericardial effusion alone).

### Differences in cardiac structure, function and arrhythmias based on troponin I status

*Table 2* demonstrates the cardiac MRI profile of the cohort which was available in 92.3% of participants. Athletes with an elevated cTnI had larger left and right ventricular chamber dimensions (p <0.001, *Figure 3*) and greater left ventricular (LV) mass compared to athletes without troponin elevation. Prevalent myocardial fibrosis was similar for athletes with cTnI elevation (30.6%) and without cTnI elevation (22.7%), (p = 0.312). There were no differences in native T1 mapping, or extracellular volume based on cTnI levels. The results were similar when evaluated based on cTnI status and antibody status (*Table S4)*.

**Figure 3.**
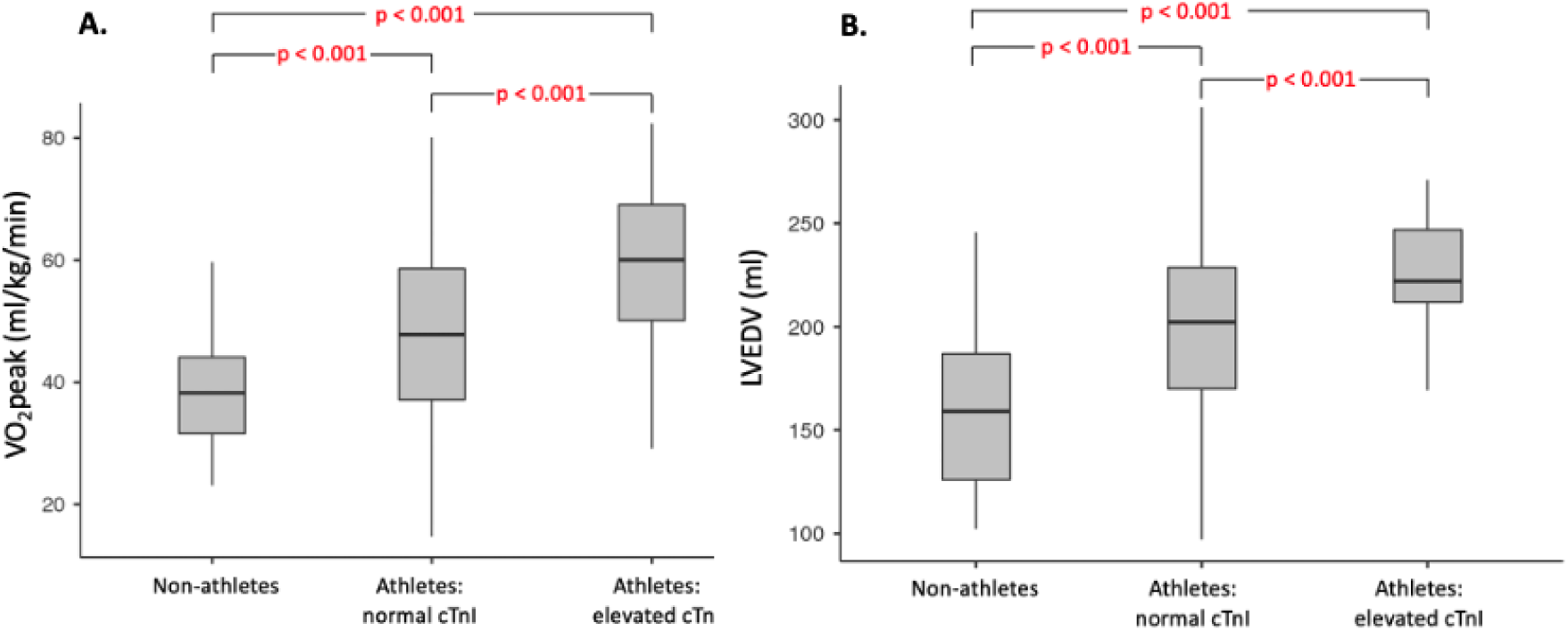
*Differences in ventricular size and fitness by troponin status.* Box plots for left ventricular end-diastolic volume (VO_2_peak; Panel A) and peak oxygen consumption (LVEDV; Panel B). The centre line denotes the median value while the grey box contains the 25^th^ to 75^th^ percentiles. P-values are considered significant if < 0.05 (red text).

**Table 2.**
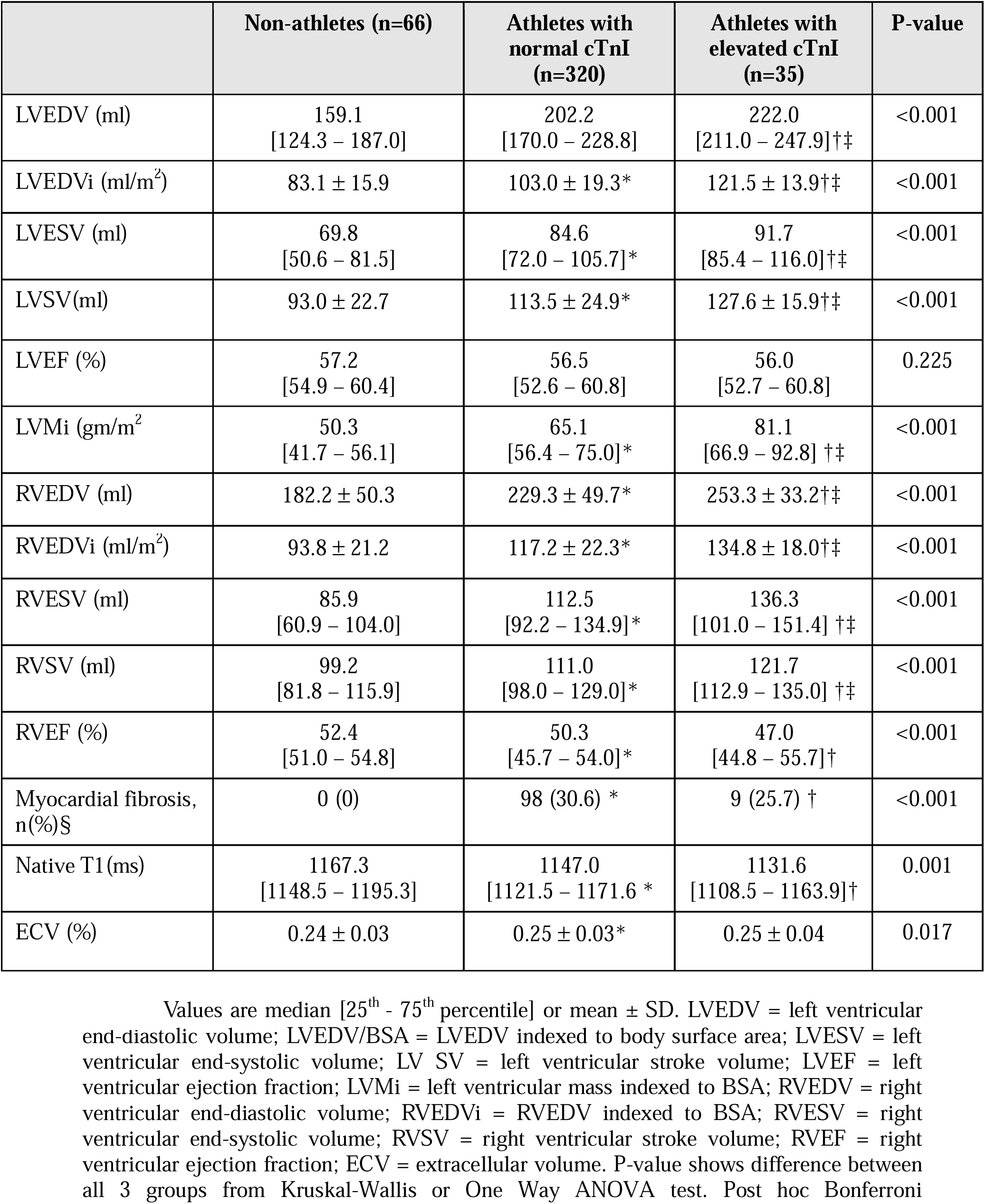
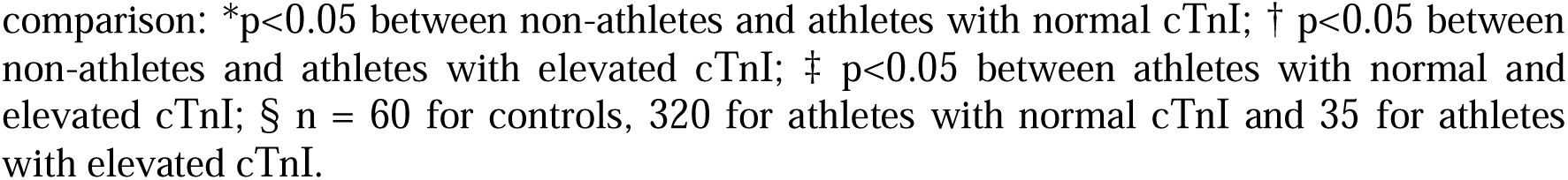
Assessment of cardiac structure, function and fibrosis with cardiac MRI by troponin status

|  | Non-athletes (n=66) | Athletes with normal cTnI (n=320) | Athletes with elevated cTnI (n=35) | P-value |
| --- | --- | --- | --- | --- |
| LVEDV (ml) | 159.1<br>[124.3 – 187.0] | 202.2<br>[170.0 – 228.8] | 222.0<br>[211.0 – 247.9]†‡ | <0.001 |
| LVEDVi (ml/m<sup>2</sup>) | 83.1 ± 15.9 | 103.0 ± 19.3* | 121.5 ± 13.9†‡ | <0.001 |
| LVESV (ml) | 69.8<br>[50.6 – 81.5] | 84.6<br>[72.0 – 105.7]* | 91.7<br>[85.4 – 116.0]†‡ | <0.001 |
| LVSV(ml) | 93.0 ± 22.7 | 113.5 ± 24.9* | 127.6 ± 15.9†‡ | <0.001 |
| LVEF (%) | 57.2<br>[54.9 – 60.4] | 56.5<br>[52.6 – 60.8] | 56.0<br>[52.7 – 60.8] | 0.225 |
| LVMi (gm/m<sup>2</sup>) | 50.3<br>[41.7 – 56.1] | 65.1<br>[56.4 – 75.0]* | 81.1<br>[66.9 – 92.8] †‡ | <0.001 |
| RVEDV (ml) | 182.2 ± 50.3 | 229.3 ± 49.7* | 253.3 ± 33.2†‡ | <0.001 |
| RVEDVi (ml/m<sup>2</sup>) | 93.8 ± 21.2 | 117.2 ± 22.3* | 134.8 ± 18.0†‡ | <0.001 |
| RVESV (ml) | 85.9<br>[60.9 – 104.0] | 112.5<br>[92.2 – 134.9]* | 136.3<br>[101.0 – 151.4] †‡ | <0.001 |
| RVSV (ml) | 99.2<br>[81.8 – 115.9] | 111.0<br>[98.0 – 129.0]* | 121.7<br>[112.9 – 135.0] †‡ | <0.001 |
| RVEF (%) | 52.4<br>[51.0 – 54.8] | 50.3<br>[45.7 – 54.0]* | 47.0<br>[44.8 – 55.7]† | <0.001 |
| Myocardial fibrosis, n(%)§ | 0 (0) | 98 (30.6) * | 9 (25.7) † | <0.001 |
| Native T1(ms) | 1167.3<br>[1148.5 – 1195.3] | 1147.0<br>[1121.5 – 1171.6] * | 1131.6<br>[1108.5 – 1163.9]† | 0.001 |
| ECV (%) | 0.24 ± 0.03 | 0.25 ± 0.03* | 0.25 ± 0.04 | 0.017 |

comparison: \* $p < 0.05$ between non-athletes and athletes with normal cTnI; † $p < 0.05$ between non-athletes and athletes with elevated cTnI; ‡ $p < 0.05$ between athletes with normal and elevated cTnI; § $n = 60$ for controls, 320 for athletes with normal cTnI and 35 for athletes with elevated cTnI.

Athletes had a higher prevalence of premature ventricular contractions (PVC) compared to controls (p < 0.001). There was no clear difference in PVC prevalence or NSVT between athletes with or without elevated cTnI or cTnI Ab. However, there was a trend towards a higher prevalence of high-burden ventricular ectopy and NSVT among athletes with elevated cTnI compared to athletes without cTnI elevation (PVC ≥ 500: 15.4% vs 7.6%, p = 0.095; NSVT: 7.7% vs 4.7%, p = 0.309, respectively) and those with cTnI Ab compared to athletes without cTnI elevation (PVC ≥ 500: 16.1% vs 7.6%, p = 0.102; NSVT: 9.7% vs 4.7%, p = 0.205, respectively)

## Discussion

In the first study to examine cTnI autoantibodies and macrotroponin in ostensibly healthy endurance athletes, we found that cTnI elevations were common, especially amongst the fittest athletes with the greatest degree of cardiac remodelling, and that these elevations were attributable to cTnI autoantibodies in a majority. This finding introduces a new paradigm for understanding troponin elevations and exercise-induced cardiac remodelling in athletes. Clinically, it challenges current practice, in which cTnI elevations in athletes are often interpreted as evidence of myocardial injury and frequently prompt comprehensive evaluation for myocardial infarction or myocarditis.

In our cohort of endurance athletes undergoing testing outside of competition, 10.1% were found to have an elevated cTnI, standing in stark contrast with the complete absence of cTnI elevations in non-athletes. These results are consistent with the few small studies which have previously assessed cTnI outside the context of intense exercise or competition^3,22^. In our study, cTnI elevations were typically modest (cTnI < 100ng/L), however two participants had cTnI values > 1000ng/L. Compared to athletes without TnI elevation, endurance athletes with cTnI measures above the normal threshold had greater cardiac remodelling (larger left and right ventricular end-diastolic volumes) and fitness (higher workload and VO_2_peak during CPET). Exercise-induced cardiac remodelling termed “the athlete’s heart” is a well-known phenomenon that is strongly correlated with fitness^23^. The extent to which troponin elevations interact with this cardiac remodelling remains controversial^6,24–26^. There is an unexplained paradox whereby troponin elevations have been accepted as a measure of myocardial injury and yet these troponin elevations were most prevalent among the highest trained individuals with measurable increases in myocardial mass. The concept that cardiac injury could promote compensatory physiological growth might seem logical, but there is little or no evidence for substantive myocyte regeneration^27,28^. Thus, it seems implausible that this damage-replacement model could explain the 150-200% increases in myocardial volumes and mass that are frequently observed among the highest trained endurance athletes^29^.

Approximately 4 out of 5 athletes with cTnI elevation were identified as having cTnI antibodies or macrotroponin (*Figure 1*). This novel finding suggests that most cases of an elevated cTnI measures in the healthy endurance athlete are not ‘true’ troponin rises. One might hypothesize that the development of anti-cardiac antibodies could occur during repeated bouts of highly strenuous exercise in which micro-damage of the myocytes could prime the immune system. The high prevalence of cTnI Ab may also be an explanation as to why previous studies have failed to demonstrate an association between exercise-induced troponin elevations and coronary atherosclerosis, and the inconsistent results reported in relation to myocardial fibrosis^4,9,30^. In our study, there was no difference in LGE between athletes with and without troponin elevation and we were unable to identify any conclusive evidence of ‘secondary’ myocardial pathology due to cTnI Ab. Conversely, it must be remembered that LGE is a relatively coarse instrument for the identification of chronic myocardial injury. We observed a trend towards a higher prevalence of high-burden ventricular ectopy and NSVT among athletes with elevated cTnI measures and suspected cTnI Ab compared to athletes without cTnI elevation. Anti-TnI Abs have previously been implicated in the development of myocardial inflammation in murine models^31,32^. However, the overall prevalence of arrhythmias among athletes in our study was too low to draw confident conclusions and it remains an open question as to whether there are any clinical sequelae associated with this potential cardiac autoimmunity and the promotion of arrhythmogenic substrate.

This study encourages clinicians to consider cTnI Ab as a cause of troponin elevation in the endurance athlete. Six athletes had a cTnI value of >200 ng/L, two of which were >1000 ng/L, all of which were due to cTnI Ab. In a clinical setting in which cTnI Ab were not considered a potential confounder, these athletes would certainly have been subjected to comprehensive evaluations, likely inclusive of invasive examinations. Thus, our research highlights the critical first step of testing for the presence of cTnI Ab when evaluating an athlete with an increase in cTnI. In our study, all endurance athletes with cTnI elevation without cTnI Ab had concomitant cTnT elevations, therefore a simple initial assessment could include testing samples with a cTnT assay followed by confirmatory testing in collaboration with local biochemistry departments. Although the co-occurrence of antibodies to cTnT and cTnI is possible, its estimated to affect <2% of healthy individuals^33^. Therefore, cTnT elevation remains an acceptable and simple way of potentially differentiating injury from antibody.

### Limitations

Although this is the first study to assess cTnI Ab in endurance athletes, there are limitations to this research. Currently, there is no gold-standard method routinely available for the direct assessment of anti-troponin antibodies. PEG leads to precipitation of high-molecular weight troponin-antibody complexes and macrotroponins. PEG precipitation methods are widely available and considered appropriate for initial assessment for the presence of antibody interference with troponin results^14^. These methods do not provide further granularity or insight into the antibody target or antigen. Finally, this cohort reflects a group of highly-trained endurance athletes outside of competition. However, despite being encouraged to refrain from vigorous exercise prior to venesection, one cannot completely exclude the possibility that the troponin elevations may have been confounded by recent bouts of exercise.

## Conclusion

Troponin I elevations are found in 10% of highly-trained endurance athletes and particularly amongst the fittest athletes with the greatest cardiac remodelling. Rather than reflecting myocardial injury, these elevations can be attributed to cTnI Ab in 80% of cases. Importantly, cTnT may aid in the assessment of true troponin elevations. These findings have immediate clinical impact. Cognizance and testing for cTnI Ab in athletes with clinically unexpected troponin elevations should help avoid intensive investigations and erroneous diagnoses.

## Data Availability

All data produced in the present study are available upon reasonable request to the authors

## Acknowledgements

None

