## Supplemental Material for "Troponin antibodies and macrotroponin: a common cause of troponin I elevation in elite endurance athletes"

**Table S1.** Cardoac Troponin I measures by sex and athlete status

|  | **Males**  **(n=334)** | | **Females**  **(n = 122)** | |
| --- | --- | --- | --- | --- |
|  | Non-athletes (n=48) | Athletes  (n=286) | Non-athletes  (n=21) | Athletes  (n=101) |
| cTnI ≥ 26 for males or ≥ 16 for females | 0 | 29 (10.1) | 0 | 10 (9.9) |
| cTnI ≥ 50 | 0 | 16 (5.6) | 0 | 5 (5.0) |
| cTnI ≥ 100 | 0 | 4 (1.4) | 0 | 2 (2.0) |
| cTnI ≥ 200 | 0 | 3 (1.1) | 0 | 1 (1.0) |
| cTnI ≥ 1000 | 0 | 1 (0.3) | 0 | 1 (1.0) |

Legend: Values are n(%). cTnI = Cardiac troponin I (ng/L)

**Table S2.** Baseline characteristics, cardiopulmonary exercise testing and Holter monitoring based on troponin and troponin antibody status

|  | **Non-athletes**  **(n=69)** | **Athletes with**  **normal cTnI**  **(n=348)** | **Athletes with**  **suspected cTnI Ab**  **(n=31)** | **P value** |
| --- | --- | --- | --- | --- |
| Age (yrs) | 23 [21 – 34] | 44 [23 – 60]* | 23 [20 – 38]‡ | <0.001 |
| Male Sex (%) | 48 (69.6) | 257 (73.8) | 21 (67.7) | 0.591 |
| BMI (kg/m^2^) | 24.3 [22.2 – 26.9] | 23.7 [22.1 – 25.5] | 21.8 [20.8 – 23.9]†‡ | 0.007 |
| Sport, n(%) | - | Athletics = 28 (8.0)  Cycling = 87 (25.0)  Rowing = 162 (46.6)  Other = 71 (20.4) | Athletics = 2 (6.5)  Cycling =16 (51.6)  Rowing = 8 (25.8)  Other = 5 (16.1) |  |
| Haemoglobin (g/L) | 146.1 ± 11.3 | 144.1 ± 9.9 | 142.2 ± 9.2 | 0.153 |
| Creatinine (µmol/L) | 72 [64 – 82] | 73 [68 -81] | 69 [65 – 74] | 0.025 |
| cTnI (ng/L) | 1 [1-2] | 3 [2-6]* | 56 [36 – 87] †‡ | <0.001 |
| cTnT (ng/L)§ | 4.4 [3.2 – 5.9] | 6.4 [4.9 – 8.9]* | 8.5 [6.4 – 11.8] †‡ |  |
| Cardiopulmonary Exercise Testing | | | | |
| Rest HR (bpm) | 75 [67 – 87] | 55 [48 – 68]* | 50 [36 – 60]† | <0.001 |
| Rest SBP (mmHg) | 134 [121 – 144] | 141 [130 – 150] | 147 [133 – 154] | 0.003 |
| Rest DBP (mmHg) | 80 [71 – 88] | 85 [77 – 94]* | 83 [69 – 91] | 0.002 |
| Peak HR (bpm) | 190 [182 – 195] | 176 [165 – 188]* | 185 [174 – 192]‡ | <0.001 |
| Peak SBP (mmHg) | 192 [172 – 212] | 213 [194 – 232]* | 218 [206 – 239]† | <0.001 |
| Peak DBP (mmHg) | 81 [75 – 88] | 85 [76 – 94]* | 82 [72 – 89] | 0.010 |
| Peak Power (W) | 266 ± 79 | 364 ± 92 * | 417 ± 75†‡ | <0.001 |
| VO_2_peak (ml/min) | 2919 [2302 – 3482] | 3669 [2847 – 4440]* | 4071 [3650 – 4853] †‡ | <0.001 |
| VO_2_peak (ml/kg/min) | 38.2 [31.6 – 44.1] | 47.8 [37.1 – 58.6]* | 59.2 [49.9 – 70.3] †‡ | <0.001 |
| VO_2_peak (% predicted) | 89.7  [76.8 – 98.0] | 126.5  [112.0 – 139.3]* | 136.1  [125.5 – 148.0]†‡ | <0.001 |
| Holter Monitoring^#^ | | | | |
| Any PVC, n (%) | 28 (41.2) | 232 (68.0) * | 19 (61.2) | <0.001 |
| PVC ≥ 100, n (%) | 6 (8.8) | 63 (18.5) * | 9 (29.0) † | 0.031 |
| PVC ≥ 500, n (%) | 1 (1.5) | 26 (7.6) * | 5 (16.1) † | 0.023 |
| NSVT, n (%) | 0 (0) | 16 (4.7) * | 3 (9.7) † | 0.041 |

Legend: Values are mean ± standard deviation, median [25^th^ - 75^th^ percentile] or n (%). BMI = body mass index; cTnI = high-sensitivity cardiac troponin I; cTnT = high-sensitivity cardiac troponin T; SBP = systolic blood pressure; DBP = diastolic blood pressure; HR = heart rate; VO_2_peak = peak oxygen consumption; W = watts; PVC = premature ventricular contraction; NSVT = non-sustained ventricular tachycardia. P-value shows difference between all 3 groups from Kruskal-Wallis or One Way ANOVA test. Post-hoc Bonferroni comparison: *p<0.05 between non-athletes and athletes with normal cTnI; † p<0.05 between non-athletes and athletes with elevated cTnI; ‡ p<0.05 between athletes with normal and elevated cTnI; § n = 58 for non-athletes, 320 for athletes with normal cTnI and 31 for athletes with elevated cTnI. # n = 68 for non-athletes, 341 for athletes with normal cTnI and 31 for athletes with elevated cTnI.

**Table S3.** PEG and cTnT results for athletes with elevated cTnI

|  | **Suspected cTnI Ab**  **(PEG recovery < 25%)**  **(n=31)** | | **True cTnI elevation**  **(PEG recovery > 25%)**  **(n=8)** |
| --- | --- | --- | --- |
|  | Normal cTnT  (n = 26) | Elevated cTnT  (n = 5) | - |
| cTnI (ng/L) | 53 [34 – 73] | 674 [474 – 1690] | 41.5 [33 – 57] |
| cTnT (ng/L) | 7.8 [6.2 – 9.6] | 26.4 [19.4 – 38.0] | 30.9 [25.1 – 45.0] |
| cTnI PEG recovery (%) | 5.0 [3.1 – 7.9] | 4.4 [4.3 – 5.7] | 46.1 [40.0 – 53.3] |
| TnT/TnI (%) | 14.7 [10.9 – 24.7] | 3.7 [2.7 – 6.3] | 81.2 [64.6 – 96.9] |

Legend: Values are median [25^th^ - 75^th^ percentile]; cTnI = cardiac troponin I when sample re-tested; PEG = polyethylene glycol; cTnT = cardiac troponin T

**Table S4.** Assessment of cardiac structure, function and fibrosis with cardiac MRI based on troponin and troponin antibody status

|  | **Non-athletes (n=66)** | **Athletes with**  **normal cTnI**  **(n=320)** | **Athletes with**  **suspected cTnI Ab**  **(n=28)** | **P-value** |
| --- | --- | --- | --- | --- |
| LVEDV (ml) | 159.1  [124.3 – 187.0] | 202.2  [170.0 – 228.8] | 231.4  [211.9 – 246.9]†‡ | <0.001 |
| LVEDVi (ml/m^2^) | 83.1 ± 15.9 | 103.0 ± 19.3* | 120.1 ± 13.6 †‡ | <0.001 |
| LVESV (ml) | 69.8  [50.3 – 81.5] | 84.6  [72.0 – 105.7]* | 98.3  [86.5 – 120.3] †‡ | <0.001 |
| LVSV(ml) | 92.5  [75.4 – 107.1] | 112.0  [96.0 – 128.5] * | 126.8  [119.2 – 132.2]†‡ | <0.001 |
| LVEF (%) | 58.1 ± 4.8 | 56.5 ± 5.9 | 55.2 ± 4.8 | 0.061 |
| LVMi (g/m^2^) | 50.3  [41.7 – 56.1] | 65.1  [56.4 – 75.0]* | 73.7 [ 64.0 -91.3]†‡ | <0.001 |
| RVEDV (ml) | 183.5 ± 48.3 | 229.8 ± 50.0* | 252.5 ± 34.6† | <0.001 |
| RVEDVi (ml/m^2^) | 93.8 ± 21.1 | 117.2 ± 22.3* | 134.7 ± 19.3†‡ | <0.001 |
| RVESV (ml) | 85.9  [60.9 – 104.0] | 112.5  [92.2 – 134.9] * | 138.2  [114.4 – 157.6] †‡ | <0.001 |
| RVSV (ml) | 99.1  [81.8 – 115.9] | 111.0  [98.0 – 129.0]* | 119.0 [110.2 – 130.8] † | <0.001 |
| RVEF (%) | 52.4  [51.0 – 54.8] | 50.3  [45.7 – 54.0]* | 45.5  [43.7 – 50.5] † | <0.001 |
| Myocardial fibrosis, n(%)§ | 0 (0) | 98 (30.6)* | 7 (25) † | <0.001 |
| Native T1(ms) | 1167.3  [1148.5 – 1195.2] | 1147.0  [1121.5 – 1171.6]* | 1130.2  [1106.1 – 1163.9] † | <0.001 |
| ECV (%) | 0.24 ± 0.03 | 0.25 ± 0.03* | 0.25 ± 0.03 | 0.012 |

Legend: Values are median [25^th^ - 75^th^ percentile] or mean ± SD. LVEDV = left ventricular end-diastolic volume; LVEDV/BSA = LVEDV indexed to body surface area; LVESV = left ventricular end-systolic volume; LV SV = left ventricular stroke volume; LVEF = left ventricular ejection fraction; LVMi = left ventricular mass indexed to BSA; RVEDV = right ventricular end-diastolic volume; RVEDVi = RVEDV indexed to BSA; RVESV = right ventricular end-systolic volume; RVSV = right ventricular stroke volume; RVEF = right ventricular ejection fraction; ECV = extracellular volume. P-value shows difference between all 3 groups from Kruskal-Wallis or One Way ANOVA test. Post hoc Bonferroni comparison: *p<0.05 between non-athletes and athletes with normal cTnI; † p<0.05 between non-athletes and athletes with elevated cTnI; ‡ p<0.05 between athletes with normal and elevated cTnI; § n = 60 for controls, 320 for athletes with normal cTnI and 28 for athletes with elevated cTnI.
